# Incidence and Clinical Presentation of Noma in Northern Nigeria (1999-2024)

**DOI:** 10.1101/2024.12.31.24319808

**Authors:** Ramat Oyebunmi Braimah, John Adeoye, Abdurrazaq Olanrewaju Taiwo, Seidu Bello, Mujtaba Bala, Azeez Butali, Bruno Oludare Ile-Ogedengbe, Abubakar Abdullahi Bello

## Abstract

Noma (Cancrum Oris), the newest neglected tropical disease, is a severe, rapidly progressing necrotizing disease of the oral cavity and facial complex with a case fatality rate of 90% if untreated. It often affects children between two and six years in Sub-Saharan Africa (Noma Belt region), with most cases reported in northern Nigeria. However, little research is available on the incidence of noma and its clinical presentation in this region using comprehensive data. Therefore, this study aims to determine the incidence of noma and its clinical presentation in northern Nigeria among different age groups. We collected retrospective data of 1,383 consecutive patients managed at Noma Children’s Hospital, Nigeria between 1999 and 2024 for incidence estimation and description of the clinical presentation of noma. Incidence calculation was done using the WHO Oral Health Unit strategy designed with the Delphi method. Our results showed that patients were between 8 months and 80 years old with a median age (IQR) of 6 years (3-15). More patients presented with acute noma than arrested noma (67.3% vs 32.7%). The estimated incidence of noma in northern Nigeria during the study period was 114.5 cases per 100,000, with Sokoto state having the highest incidence of 921.9 cases per 100,000, while Adamawa state had the lowest incidence of 6.3 cases per 100,000. The annual average and median incidence of noma across all years was 4.4 and 2.2 cases per 100,000 (range: 0.2-21.3 cases per 100,000), although between 2020 and 2024, the annual average and median incidence estimates were 15.5 and 15.6 cases per 100,000. Also, this study found the incidence of noma cases with gangrene to be higher than cases with oedema or acute necrotizing ulcerative gingivitis. These findings confirm the high incidence and impact of noma in northern Nigeria in the last two and half decades and highlight the need to intensify awareness of risk factors and early signs of noma within communities in the region and to conduct community-based screening to promote the identification and cost-effective treatment of reversible early noma disease.

**Author Summary:** Noma is a severe and deadly gangrenous disease that affects the mouth and other tissues of the face. It is often associated with poverty and is currently prevalent in Sub-Saharan Africa. More cases have been described in northern Nigeria than in any African region. However, the incidence of noma and its clinical presentation in this region is unknown from data available from different states. In this study, we retrospectively collected data from the only dedicated specialist hospital for noma intervention in Nigeria between 1999 and 2024 to calculate noma incidence within this period. Then the incidence of noma was calculated using a method proposed by the World Health Organization (WHO). This study estimates an incidence of 114.5 cases per 100,000 within the study period and an annual average and median incidence of 4.4 and 2.2 cases per 100,000. Of note, the incidence of noma with gangrene (stage 3) was higher than the incidence of oedema (stage 2) or acute necrotizing ulcerative gingivitis (stage 1). These results highlight the importance and impact of noma in northern Nigeria and underscore the need for noma awareness and screening programs for population education and detection of early noma disease which are reversible with cost-effective interventions.

## Introduction

Noma is a devastating and disfiguring necrotizing disease that primarily affects malnourished children in low-income countries. With a reported mortality rate exceeding 90% in untreated cases, noma has been recognized as a public health challenge for many countries in sub-Saharan Africa ^1^. Noma is most prevalent in the areas mapped out as the Noma belt spanning far east from Mauritania, Senegal, Mali, Niger, Chad, Sudan, and Ethiopia in far West ^2,3^. This disease predominantly affects children between the ages of two and six years old in poorly developed countries where adequate nutrition, sanitation, and cleanliness are lacking ^1^. On the contrary, acute noma cases have been reported in northern Nigeria in adult patients ^4^. Despite its severe impact, the aetiology of noma remains poorly understood, which limits effective preventive and therapeutic strategies ^5^. Historically, noma was believed to be associated with a combination of malnutrition, poor oral hygiene, measles, and bacterial infections ^5,6^. However, recent evidence suggests a multifactorial origin involving complex interactions between genetic predisposition, host immune response, environmental factors, and microbial dysbiosis ^7–9^.

The geographical distribution and prevalence of noma continue to generate major questions globally because very little data exists about its incidence, mortality rate, and other epidemiological factors. Similarly, reasons for noticeable dissimilarities in the incidence and prevalence among comparable population groups of different but equally impoverished and underdeveloped countries remain vague ^10,11^. To emphasize research that measure the efficacy of any preventive intervention by policymakers, it is important to record and properly understand the epidemiology of the disease condition ^12^. Therefore, this present study aimed to determine the incidence and clinical presentation of noma in northern Nigeria which falls within the “noma belt” of sub-Saharan Africa.

## Methods

This retrospective study involves the health records of consecutive patients with noma managed at the Noma Children’s Hospital (NCH), Sokoto Nigeria, from September 1999 to October 2024. Founded in 1999, NCH is the only specialized health facility in Nigeria, and one of few worldwide, dedicated to managing acute noma and noma survivors. The model of care at the hospital involves the provision of intensive care during active disease, management of noma sequelae, multidisciplinary care for patients with noma, and community-based services ^13^.

### Data Collection

Eligible patients for data extraction from medical records were those diagnosed with noma (active disease and post-disease defects) at NCH within the study period who resided in any of the nineteen northern Nigerian states (Adamawa, Bauchi, Benue, Borno, Gombe, Jigawa, Kaduna, Kano, Katsina, Kebbi, Kogi, Kwara, Nasarawa, Niger, Plateau, Sokoto, Taraba, Yobe, Zamfara) and Federal Capital Territory at the time of diagnosis The rationale for limiting data collection to Northern Nigeria was due to the predominant number of noma cases from this region ^14–16^ and NCH’s location in the country. Individuals across all age groups were included. However, patients with necrosis or gangrene affecting anatomic sites outside the orofacial region were not considered. Also, we excluded records of patients who resided outside Nigeria at the time of diagnosis, and patients with missing data on their state of residence at diagnosis were excluded. Patient information was collected using an electronic spreadsheet, and variables collected included demographic data (i.e., age and sex), year of diagnosis, state of origin, and state of residence at diagnosis. Also, the orofacial sites affected by noma, body weight at diagnosis, and hemoglobin concentration were collected. Patients were also categorized according to the five stages of disease progression recommended by the World Health Organization (WHO) i.e., acute necrotizing ulcerative gingivitis (ANUG; 1), oedema (2), gangrene (3), scarring (4), and sequelae (5). In this study, “acute noma” refers to noma stages 1 to 3 while “arrested noma” is used as a collective term for noma stages 4 and 5.

### Statistical Analysis

Data was analyzed using Statistical Package for the Social Sciences (SPSS) v 29 (IBM Corp, Armonk, NY, USA). Descriptive analysis was performed for categorical and continuous variables and presented as texts, tables, and figures. The normal distribution of continuous variables was also determined using Kolmogorov-Smirnov’s test before significant differences (or otherwise) by categories were determined using Mann-Whitney U test (for two categories) or Kruskal-Wallis test (for three or more categories). Pearson’s Chi-square test was conducted to determine significant differences in the proportion of categorical variables if relevant statistical assumptions were met. Else, Fisher’s exact test was used. For all tests, probability values below 5% were considered statistically significant.

This study estimated the incidence of noma in the study period considering only patients with acute disease. Incidence analysis was done following the WHO Oral Health Unit’s 1994 expert consultation report conducted using the Delphi method as a two-step process ^14,17,18^. Also, only data from patients with acute noma (WHO stages 1 to 3) were used for incidence calculation. Two states (Nasarawa and Plateau) had no cases with acute noma in the dataset and were excluded from further analysis. First, the total number of surviving cases (*S*) was determined as a ratio of the number of cases referred to NCH (provided by our data, *R*) and the estimated percentage of surviving cases that were referred (*x*). This is given by the equation:

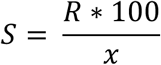

Where *x* was approximately 15% based on previous reports and experts’ opinions ^14,17^. Then, the incidence of noma (*I*) was calculated as the ratio of the number of surviving cases (*S*) and the case survival rate of noma (*y*) which was 10% according to previous studies ^17,19,20^.

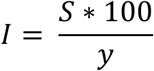

The incidence of noma was also determined for each age category, sex, and state of residence at diagnosis. The 2006 Nigerian population census results served as the reference population used for reporting incidence values in this study ^21^. Also, the total population for the states represented in the observed number of cases in our dataset (numerator) was considered during the overall incidence analysis. Due to the hospital-based dataset employed, this study did not perform prevalence analysis since NCH data only accounts for a fraction of the expected total cases (especially for patients with post-noma defects) that were not considered during prevalence estimation.

### Ethics

Approval to conduct this study was granted by the Sokoto State Ministry of Health Ethics Research Committee (Registration no: SKHREC/037/2024). Informed consent was waived due to the retrospective nature of this study. All potential patient identifiers were removed from the data collected, and anonymized data was used for analyses.

## Results

### Patient Demographics

One thousand three hundred and eighty-three patients with noma managed at NCH between 1999 and 2024 were included for analysis in this study. Detailed description of patients and their demographic characteristics are presented in Table 1. Patients were between 8 months and 80 years old with a median age (IQR) of 6 years (3-15). More patients were below 5 years old (n=569, 44.1%), while the proportions of children between five and nine years old, adolescents (10-17 years), young adults (18-39 years), middle-aged adults (40-64 years), and elderly patients (≥65 years) were 24.5% (n=339), 11.9% (n=165), 16.3% (n=225), 5.2% (n=72), and 0.9% (n=13) respectively. Also, most patients with noma were males (n=750, 54.2%) than females (n=633, 45.8%). According to the state of residence, most patients were from Sokoto state (n=709, 51.3%), followed by Zamfara (n=173, 12.5%), Kebbi (n=170, 12.3%), and Kano state (n=137, 9.9%). More noma cases were observed in 2021 (n=220, 15.9%) than in other years, with patients between 2021 and 2024 accounting for 51.7% of all cases (n=714) (Figure 1). When the year of diagnosis was stratified by the state of residence, the analysis showed that Sokoto state had the highest number of cases in 23 of 26 years considered in this study (range: 3 cases in 2009 to 116 cases in 2021; p<0.001) (Figure 2). Kano state had the highest number of cases in 2000, while Kebbi state had the highest number in 2019.

**Figure 1:**
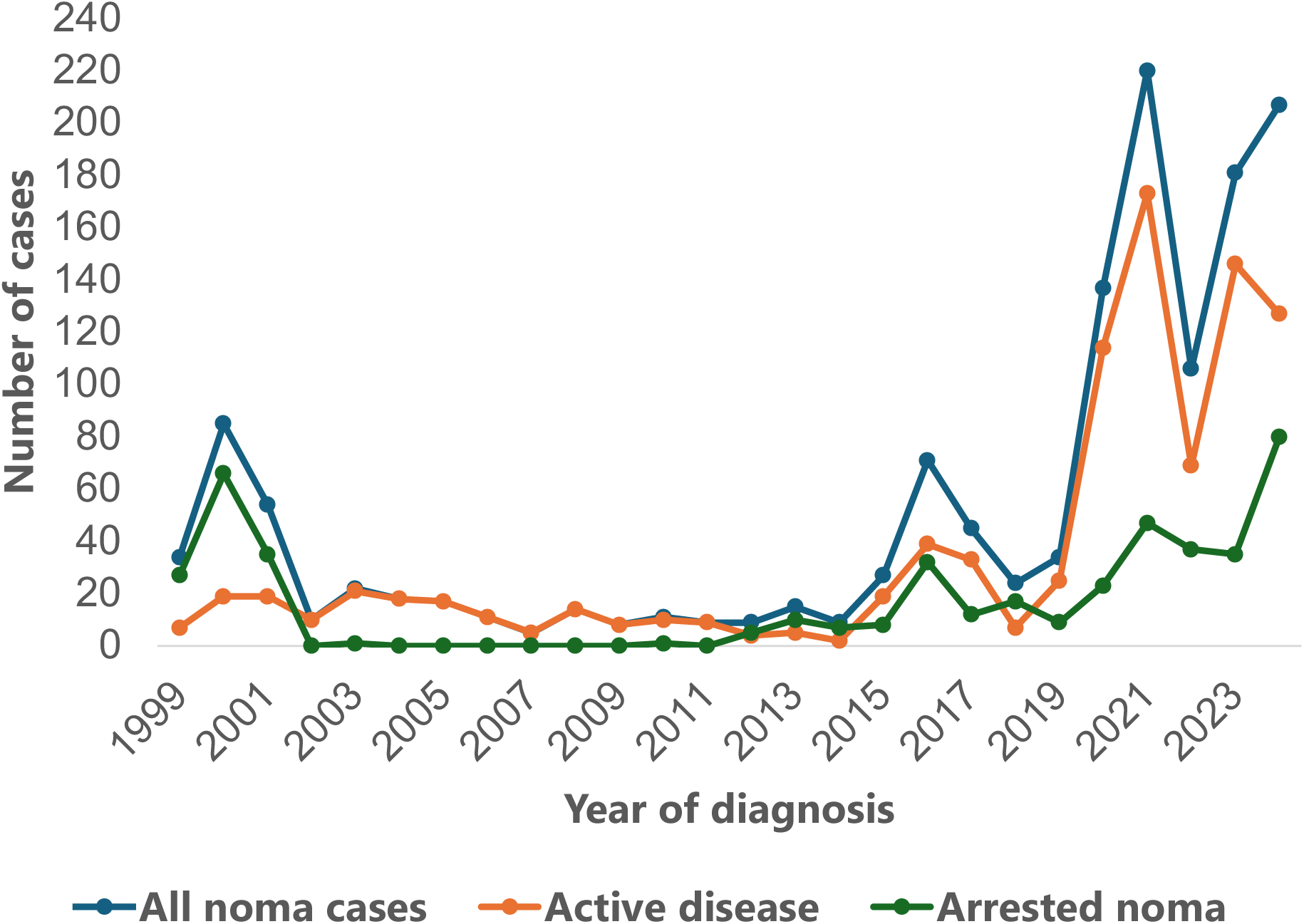
Line plot showing the annual distribution of noma cases between 1999 and 2004.

**Figure 2:**
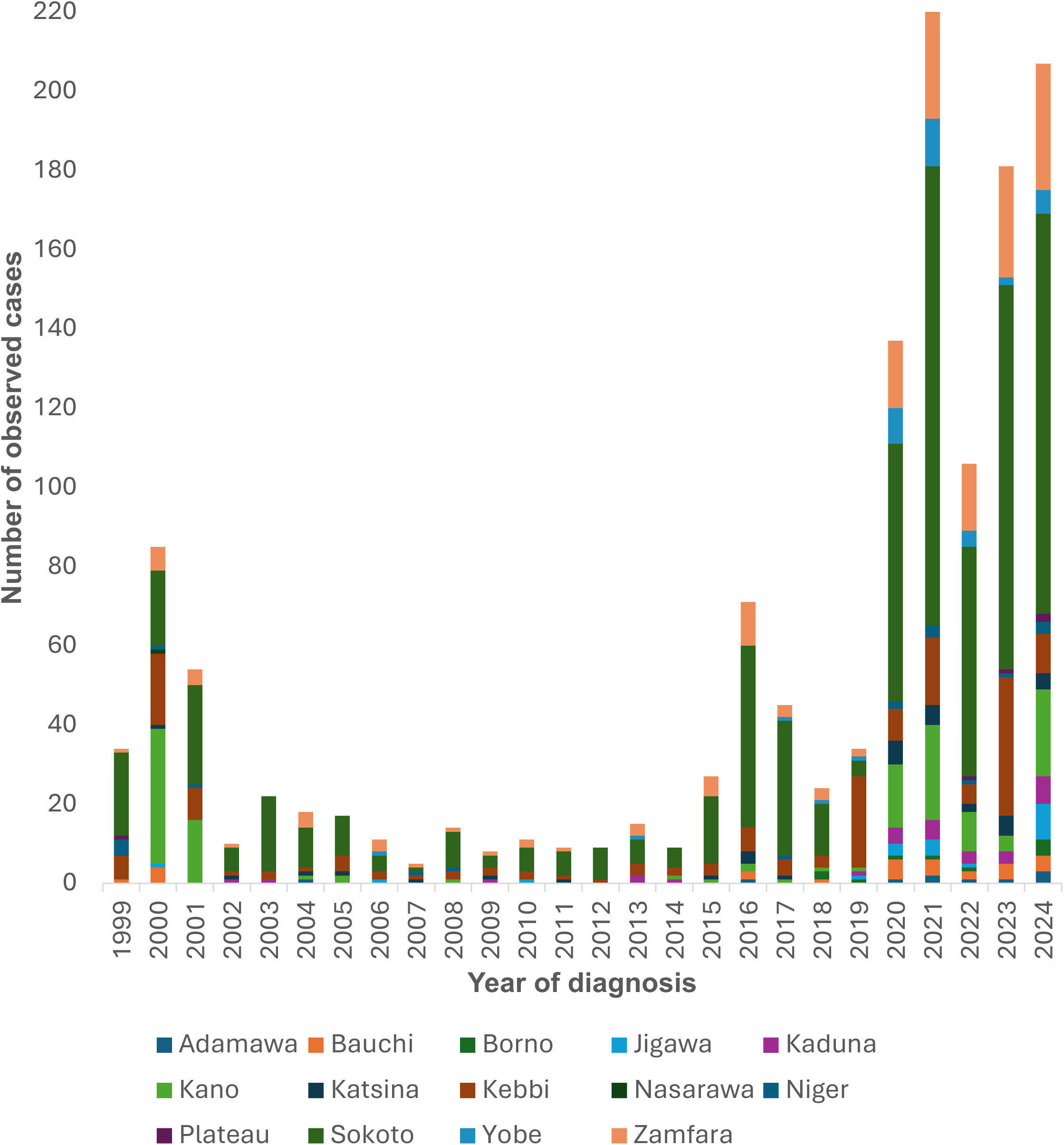
Stacked bar plot showing the number of noma cases between 1999 and 2024 by their state of residence at diagnosis

**Table 1:**
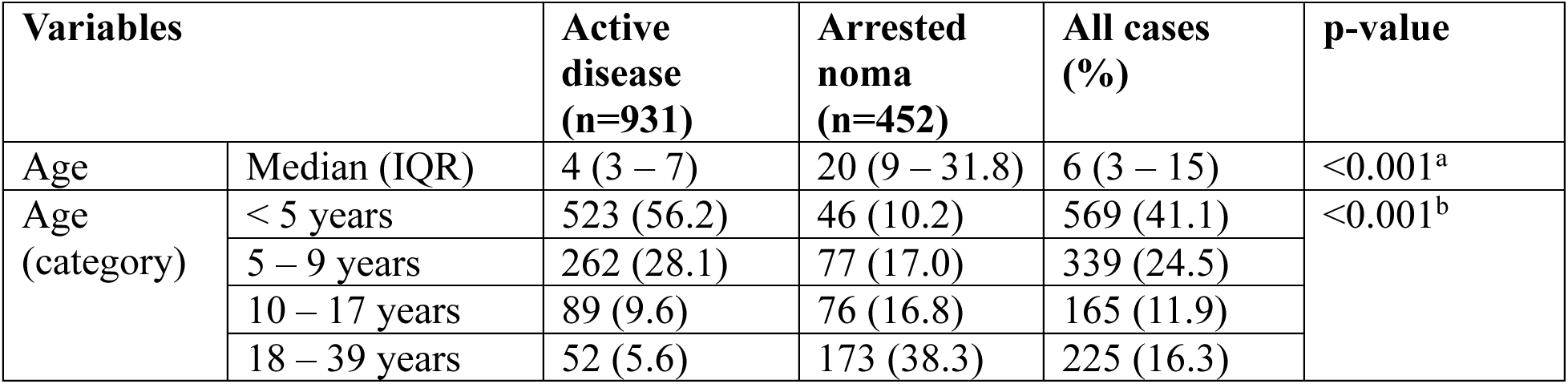
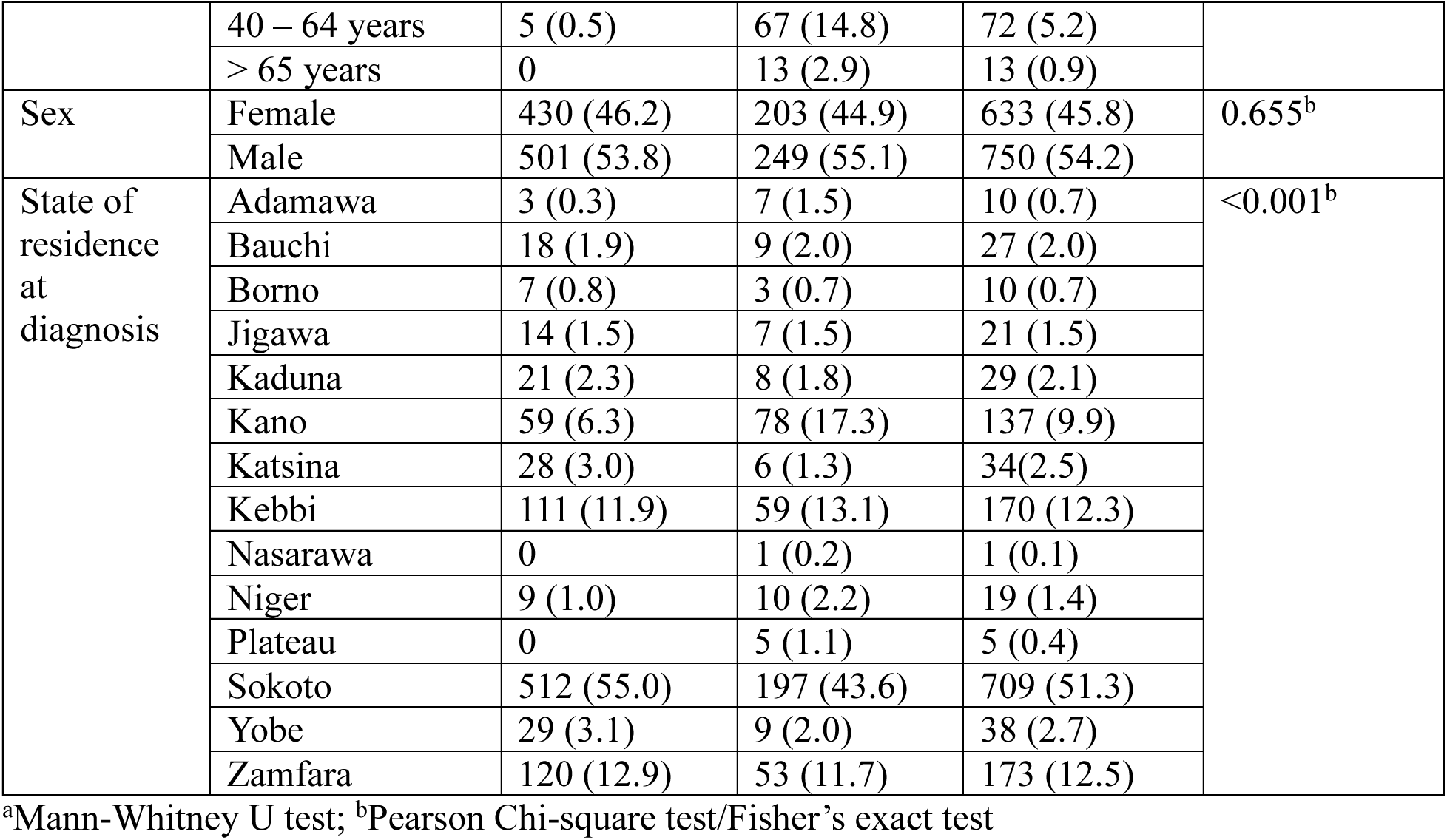
Demographic distribution of 1383 noma cases managed between 1999 to 2024.

### WHO Staging and Clinical Presentation

More individuals presented with acute noma than arrested noma (n=931, 67.3% vs n=452, 32.7%) (Table 1). Upon stratifying the active noma disease by the three pertinent WHO stages (i.e., stages 1 to 3), the analysis showed that most patients had gangrene (n=825, 59.7%) than oedema (n=101, 7.3%), or ANUG (n=5, 0.4%). Likewise, for arrested noma (i.e., stages 4 and 5), more patients have scarring (n=346, 25.0) than sequelae of acute noma (n=106, 7.7%) (Table 2). Cases with arrested noma had a significantly higher median age than cases with active disease (20 years vs 4 years old, p<0.001). Following age stratification, this study also found that a significantly higher proportion of patients with active disease were below five years old (n=523, 56.2%) while all patients that were 65 years and above had arrested noma (n=13, 2.9%) (p<0.001). A detailed analysis of the patient’s age and the five noma stages is also shown in Table 2. However, a similar proportion of males and females had acute and arrested noma (p=0.655, Table 1). This study also found that a significantly higher proportion of cases from Sokoto state were acute noma (55% vs 43.6%), while more cases with arrested noma were from Kano (17.3% vs 6.3%) and Kebbi states (13.1% vs 11.9%) (p<0.001, Table 1).

**Table 2:** Clinical stage and presentation of 1383 noma cases managed between 1999 to 2024.

| Variables |  | Age of patients |  |  |  |  |  | All cases | p-value |
| --- | --- | --- | --- | --- | --- | --- | --- | --- | --- |
|  |  | <5 years | 5–9 years | 10–17 years | 18–39 years | 40–64 years | >65 years |  |  |
| WHO noma stage ($n=1383$) | 1 | 3 (0.5) | 2 (0.6) | 0 | 0 | 0 | 0 | 5 (0.4) | <0.001<sup>a</sup> |
|  | 2 | 81 (14.2) | 11 (3.2) | 6 (3.6) | 3 (1.3) | 0 | 0 | 101 (7.3) |  |
|  | 3 | 439 (77.2) | 249 (73.5) | 83 (50.3) | 49 (21.8) | 5 (6.9) | 0 | 825 (59.7) |  |
|  | 4 | 42 (7.4) | 72 (21.2) | 73 (44.2) | 124 (55.1) | 28 (38.9) | 7 (53.8) | 346 (25.0) |  |
|  | 5 | 4 (0.7) | 5 (1.5) | 3 (1.8) | 49 (21.8) | 39 (54.2) | 6 (46.2) | 106 (7.7) |  |
| Body weight (kg) (n=858) | Median (IQR) | 8.8 (7.1-10.6) | 15 (11.1-19) | 31 (20-38.3) | 52 (45-60) | 55.5 (48-62.3) | 42.5 (36-67) | 15 (9.3 – 41) | <0.001<sup>b</sup> |
| Hemoglobin concentration (g/dL) (n=921) | Median (IQR) | 9.3 (7.4-11.2) | 11 (9.6-12.1) | 12.1 (10 – 13.1) | 13.1 (11.4-14.8) | 12.2 (11.5 – 14) | 7.8 (7.8-9.5) | 11.1 (9.1 – 12.6) | <0.001<sup>b</sup> |
| Anatomic sites affected (n=1383) | Nose | 15 (2.6) | 3 (0.9) | 0 | 0 | 0 | 0 | 18 (1.3) | 0.012<sup>a</sup> |
|  | Right cheek | 272 (47.8) | 187 (55.2) | 100 (60.6) | 154 (68.4) | 46 (63.9) | 9 (69.2) | 768 (55.5) | 0.012<sup>a</sup> |
|  | Left cheek | 283 (49.7) | 167 (49.3) | 89 (53.9) | 106 (47.1) | 34 (47.2) | 4 (30.8) | 683 (49.4) | 0.578<sup>a</sup> |
|  | Upper lip | 17 (3.0) | 0 | 0 | 0 | 0 | 0 | 17 (1.2) | <0.001<sup>a</sup> |
|  | Lower lip | 21 (3.7) | 4 (1.2) | 0 | 0 | 0 | 0 | 25 (1.8) | <0.001<sup>a</sup> |
|  | Eye | 7 (1.2) | 3 (0.9) | 2 (1.2) | 5 (2.2) | 1 (1.4) | 0 | 18 (1.3) | 0.828<sup>a</sup> |

<sup>a</sup>Pearson's Chi-square test/Fisher's exact test; <sup>b</sup>Kruskal-Wallis test

The median body weight (IQR) of all noma cases at diagnosis was 15 kg (9.3-41), and the detailed stratification of the median body weights by different age groups is shown in Table 2. This ranged from 8.8 kg (7.1-10.6) among children below five years to 55.5 kg (48-62.3) among cases who were between 40 and 64 years old (i.e., middle age group). The median hemoglobin concentration (IQR) at diagnosis for all cases was 11.1 g/dL (9.1-12.6), ranging from 7.8 g/dL (7.8 – 9.5) among elderly cases (above 65 years old) to 13.1 g/dL (11.4-14.8) among young adults (18 to 39 years old) (Table 2).

Analysis based on the sites affected by noma showed that the right cheek was affected in 55.5% (n=768) of patients, while the left cheek was affected in 49.4% of patients (n=683). The lower lip was involved in 1.8% of patients (n=25) while 1.3% (n=18) of patients had noma disease that affected the nose and eyes. Also, upper lip involvement was observed among 1.2% of patients (n=17). Of note, significantly more patients below five years old had diseases that affected the nose and both lips while more patients who had noma that affected the right cheek were above ten years old (p<0.001-0.012).

### Incidence of Noma

The estimated total number of new cases of noma in northern Nigeria between 1999 and 2024 was 62,067, with an overall incidence of 114.5 cases per 100,000 population. The overall incidence among males was 120.7 cases per 100,000 population, while 108.1 cases per 100,000 population was calculated for females. Figure 3 shows the incidence of noma within the study period, with Sokoto state having the highest incidence of 921.9 cases per 100,000 population and Adamawa state having the lowest incidence of 6.3 cases per 100,000 population. Within the study period, the annual incidence of noma ranged from 0.2 cases per 100,000 population in 2014 to 21.3 cases per 100,000 in 2021 (Figure 4). Notably, this study observed that the incidence of noma increased significantly between 2020 and 2024 (8.5 cases per 100,000 population to 21.3 cases per 100,000 population) compared to other periods (Figure 4). The average and median incidence values across all years was 4.4 and 2.2 cases per 100,000, while the average and median incidence between 2020 and 2024 was 15.5 and 15.6 cases per 100,000 respectively.

**Figure 3:**
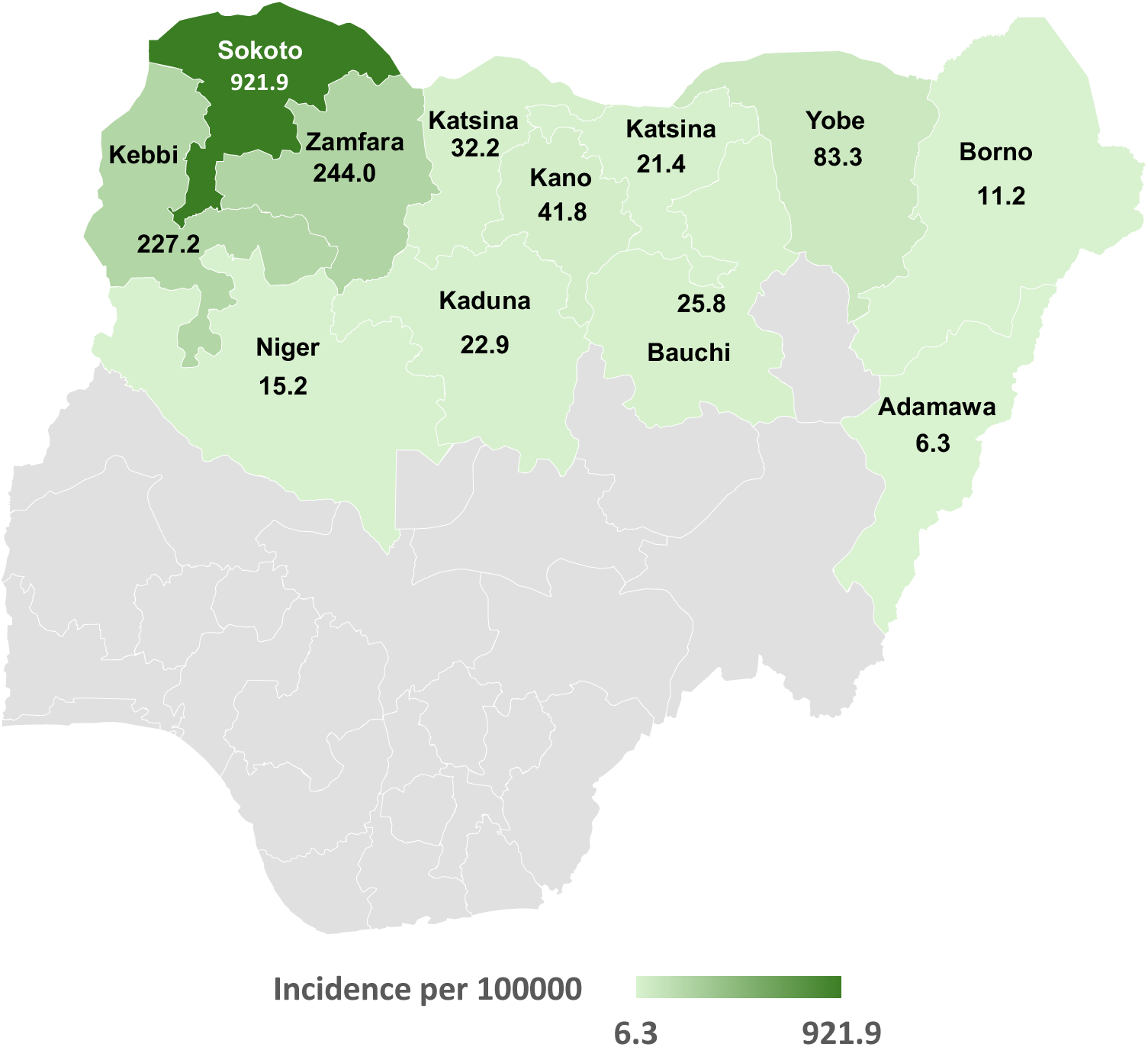
Map of Nigeria showing the estimated incidence of noma (per 100,000 population) in different states in Northern Nigeria from 1999 to 2024.

**Figure 4:**
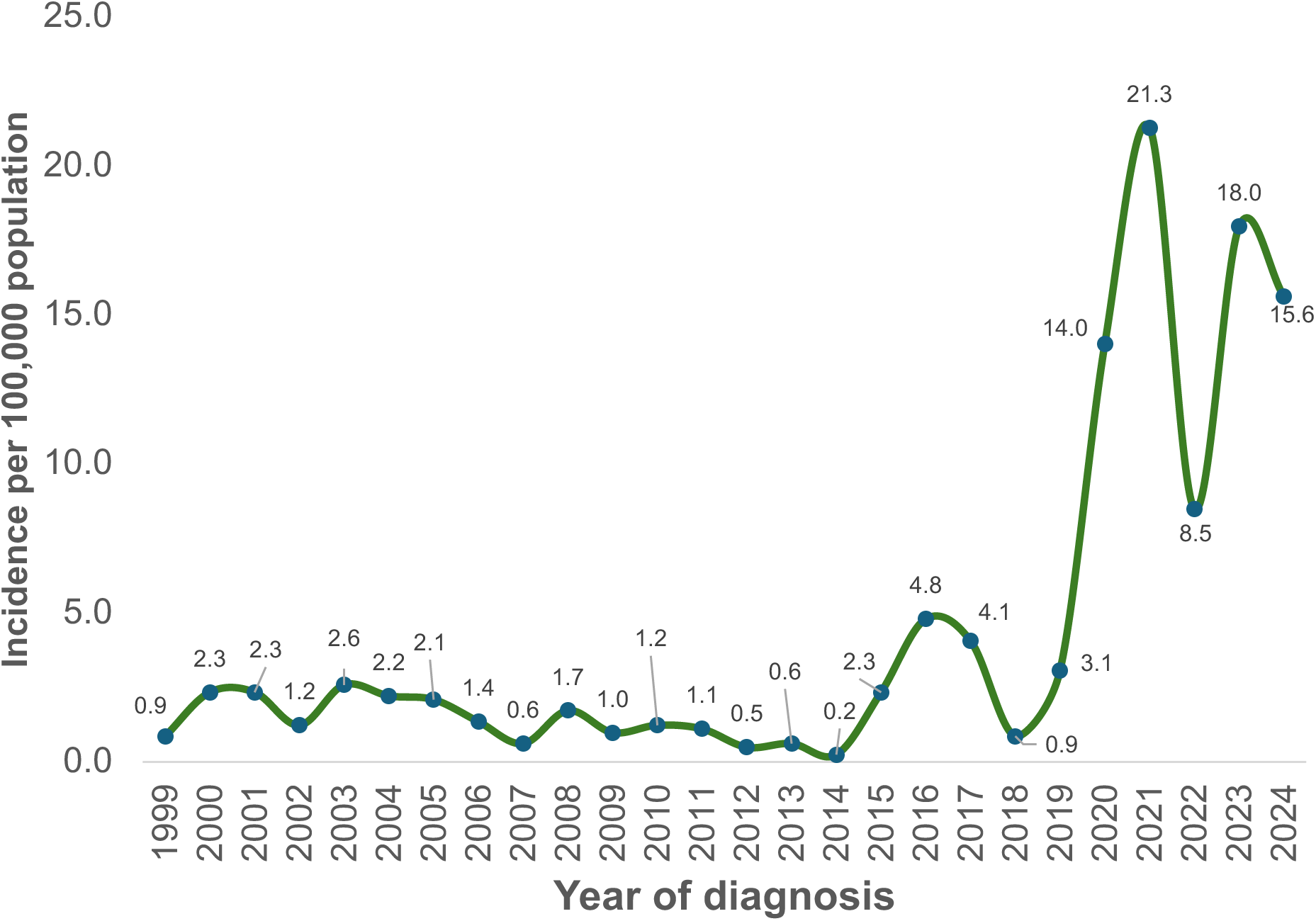
Noma incidence in Northern Nigeria by year of diagnosis from 1999 to 2024

When stratified by the WHO noma stages 1 to 3, this study found the incidence of ANUG to be 0.6 cases per 100,000 population, 12.4 cases per 100,000 population for patients with oedema, and 101.5 cases per 100,000 population for patients with gangrene. Compared to other age groups, noma incidence was higher among patients below ten years old, i.e., 399.9 cases per 100,000 among children below five years and 226.2 cases per 100,000 population among children between five and nine years old (Figure 5). Among individuals between 0 and 17 years, the incidence of noma was higher in Sokoto state than in other states within Northern Nigeria (382.9 to 3827 cases per 100,000 population, Figure 6). However, noma incidence among young adults was higher in Kebbi state (128.7 cases per 100,000 population) compared to other states studied. Also, new noma cases among middle-aged adults were only recorded in Kebbi state within the study period (Incidence: 72.5 cases per 100,000 population, Figure 6).

**Figure 5:**
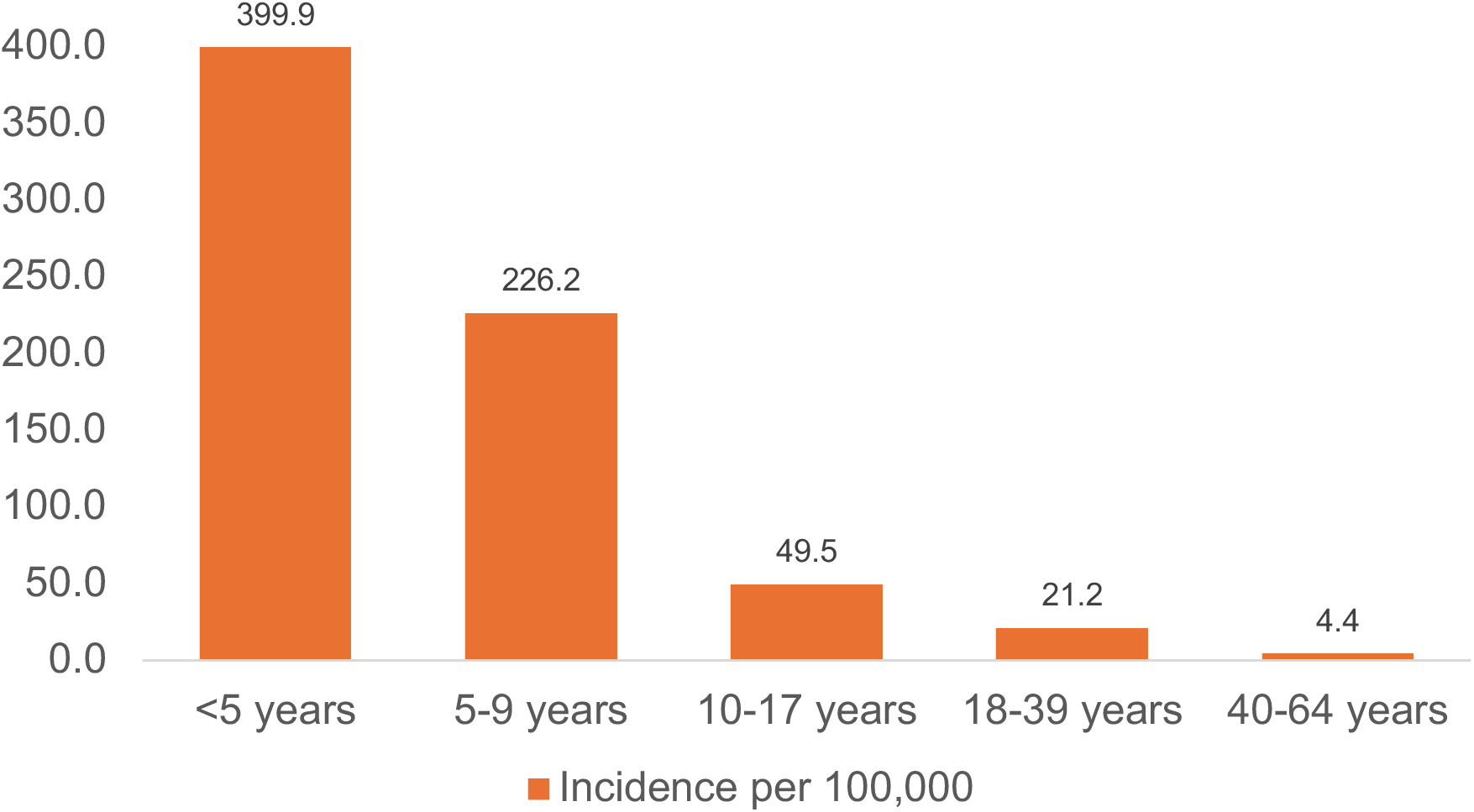
Bar plot showing noma incidence among different age groups in Northern Nigeria.

**Figure 6:**
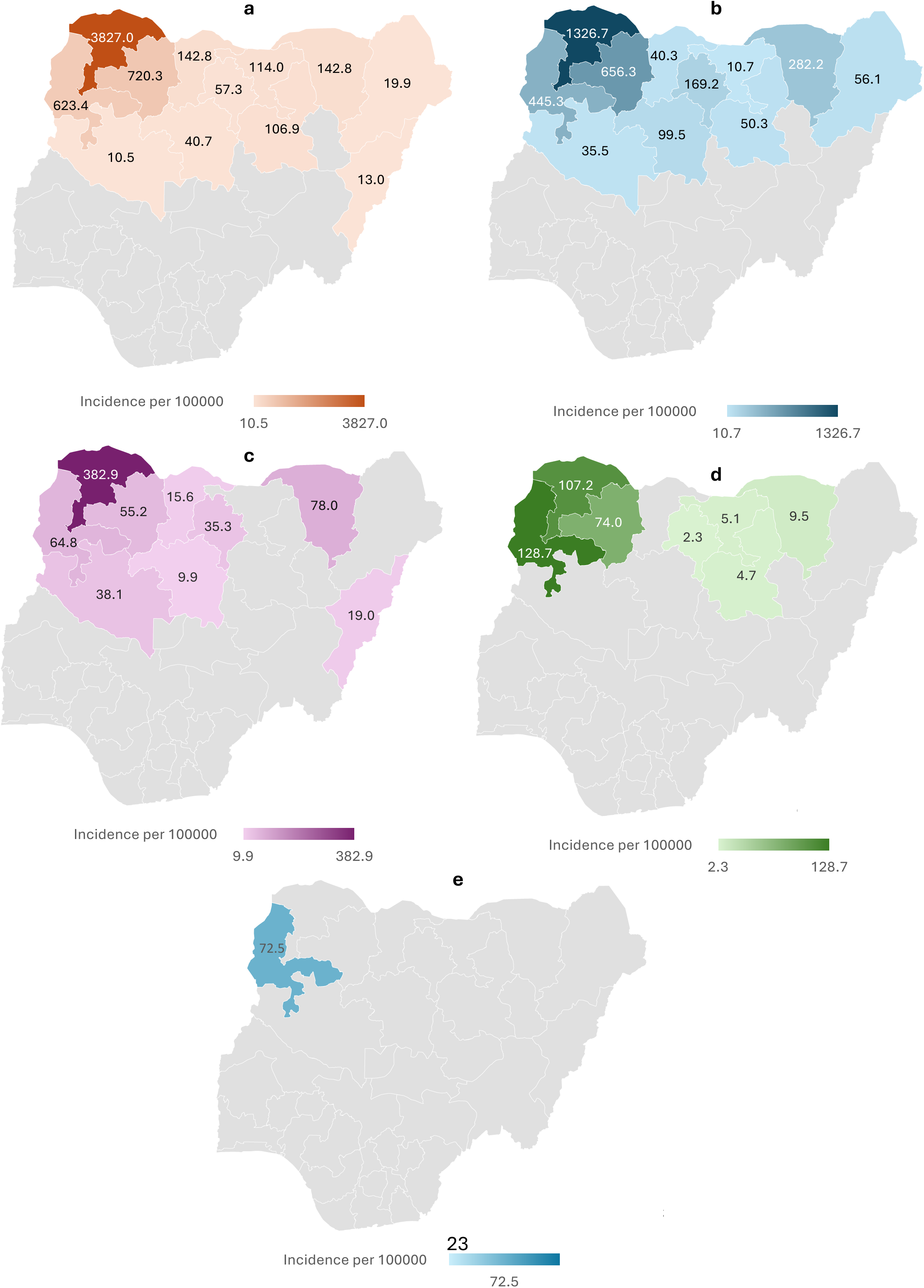
Map showing the incidence of noma (per 100,000) population during the study period among different age groups (a) < 5 years (b) 5 – 9 years (c) 10 – 17 years (d) 18 – 39 years and (e) 40 – 64 years

## Discussion

Noma, the most recent addition to the WHO’s Neglected Tropical Diseases list, is a severe necrotizing disease of the orofacial complex with a case fatality rate of 90%. It often affects children between two and six years within Sub-Saharan Africa (Noma Belt region), and most cases from this region in the last three decades have been reported in northern Nigeria ^16,22^. Given the scarcity of population data on noma incidence due to its association with low-income populations, this study employs data from a large specialist hospital dedicated to noma prevention and management to estimate the incidence of noma and determine its clinical presentation in northern Nigeria. This is the first study to calculate the incidence of noma and describe its clinical presentation across different states within northern Nigeria. Our analysis estimated the incidence of noma in northern Nigeria, from 1999 to 2024, is 114.5 cases per 100,000, with a slightly higher incidence of 120.7 cases per 100,000 among males and 108.1 cases per 100,000 among females living in the region. Moreover, the incidence of noma was found to vary between 0.2-21.3 cases per 100,000 annually, with Sokoto and Adamawa states having the most and least incidence respectively. This study also found that the overall incidence of noma was generally higher among patients below 10 years old. However, noma incidence among persons between 18 and 64 years old was more common in Kebbi state compared to other areas within northern Nigeria.

The incidence of noma in the northern Nigeria subregions has been investigated in different studies ^14,22,23^. Recently, using twelve acute noma cases encountered between 2010 and 2018, Bello et al estimated the incidence of noma within north-central Nigeria to be 8.3 cases per 100,000 population, ranging from 4.1 to 17.9 cases per 100,000 in the different states that make up the subregion ^14^. In this study, we estimated that the incidence of noma in northern Nigeria is 114.5 cases per 100,000 population, which comparatively, is about fourteenfold higher than the incidence estimated for north-central Nigeria alone. Though similar methods were used to estimate the incidence of noma in Bello et al’s study and ours, this finding may be attributed to the wider regional scope and study period considered in this study. In Sokoto state (northwest Nigeria), Fieger et al also analyzed 378 noma cases and estimated an average incidence of 640 cases per 100,000 among persons aged 10 to 30 between 1996 and 2001 (range: 440-850 cases per 100,000 population) ^23^. In this study, the incidence of noma among persons between 10 and 40 years in Sokoto ranged between 107.2 and 382.9 cases per 100,000, which is lower than the estimates reported by Fieger et al. This finding may be related to the different approaches for incidence estimation in both studies, with Fieger et al calculating noma incidence using the incidence of cleft lip, age, location of the patients relative to the treatment center, and noma mortality. Further comparing our calculated incidence with those reported by Lafferty between 2002 and 2012 ^24^, this study finds that the incidence of noma in northern Nigeria is about twenty folds higher than reported incidences of noma in Ethiopia among children between 0 and 9 years old.

This study found that the incidence of noma was higher in Sokoto than in other states within northern Nigeria. This finding supports the systematic review by Galli and colleagues ^22^, highlighting that Sokoto was the subnational region with the highest number of new noma cases in Africa. This finding is expected given that Sokoto state has the highest poverty rate in Nigeria^25^. However, our analysis showed that this notion holds only for patients below 18 years, but not young and middle-aged adults with acute noma, where the highest incidence during the study period was observed in Kebbi state. Noma in adults is usually associated with immunocompromising conditions like HIV infection/AIDS and leukemia, and this study’s finding may be related to the higher HIV prevalence in Kebbi than in Sokoto state ^26^.

Our findings on the pattern of acute noma presentation and staging of noma in northern Nigeria showed that there were significantly more patients with gangrene (irreversible stage) than oedema or ANUG (reversible stages) during the study period. This further supports that noma presentation in northern Nigeria is frequently in the later stages of the disease and highlights the need to intensify efforts aimed at creating awareness of the early signs of noma in the region and screening children below ten years to identify those with simple gingivitis, ANUG, and oedema for referral and management. In this study, noma mostly involved the cheeks, followed by the lower lip, nose, and eyes. This finding is similar to those of Adeniyi and Awosan ^27^, who used a subgroup of the data employed in this study to describe the pattern of noma in northwest Nigeria. However, Bello et al ^14^ found that noma in northcentral Nigeria often affected the nose and upper lip than other sites, which is in contrast to our observation on the affected sites.

Though our study uniquely estimated noma incidence in northern Nigeria with the largest cohort of noma cases to date, it is not without limitations. The main limitation of this study is the lack of data from six out of 20 eligible northern Nigeria states/territories, i.e., Federal Capital Territory, Gombe, Taraba, Benue, Kwara, and Kogi. However, the latter three states have the lowest poverty rates in northern Nigeria, which may indicate a low incidence/prevalence of noma in these areas. Little is known about the number of noma cases from Gombe or Taraba to date, which remains to be explored in future studies. Also, given that a retrospective cohort was used for analysis, this study did not examine the detailed risk factors of noma in the different areas.

## Conclusions

This study estimated the incidence of noma in northern Nigeria between 1999 and 2024 to be 114.5 cases per 100,000 population, with a slightly higher incidence among males than females. The annual average and median incidence of noma across all years was 4.4 and 2.2 cases per 100,000 (range: 0.2-21.3 cases per 100,000). Notably, Sokoto, Kebbi, and Zamfara states had the highest incidence estimates, while Adamawa state had the lowest incidence in the study period. However, this study found that the association between incidence and state of residence was related to the individual’s age, with Sokoto state having a high incidence of noma cases among those below 18 years, while Kebbi state had the highest incidence of noma cases among those between young and middle-aged adults. Noma incidence was highest among populations below ten years old than other age groups. Also, the incidence of cases with gangrene was significantly higher than cases with oedema or ANUG.

## Data Availability

Data used for this study is not publicly available due to the need to maintain patient confidentiality. However, anonymized data may be obtained from the corresponding authors upon reasonable request.

